# Mid-pregnancy sleep disturbances are not associated with mid-pregnancy maternal glycemia

**DOI:** 10.1101/2023.04.21.23288767

**Authors:** Marquis Hawkins, Maisa Feghali, Kaleab Z Abebe, Christina M. Scifres, Christina M Lalama, Tina Costacou, Patrick Catalano, Hyagriv Simhan, Steve Orris, Dara Mendez, Daniel J. Buysse, Esa M Davis

## Abstract

**Background:** In pregnancy, epidemiological data have consistently shown strong associations between sleep quality and duration and maternal glycemia. However, other sleep disturbances such as difficulty falling asleep and staying asleep are common in pregnancy. They may contribute to impaired maternal glycemia through sympathetic nervous system activity, systemic inflammation, and hormonal pathways. However, there is little research examining associations between these specific sleep disturbances and maternal glycemia.

**Objective:** This study aimed to investigate the associations of sleep disturbances during mid-pregnancy and mid-pregnancy maternal glycemia and gestational diabetes subtypes.

**Study Design:** This is a secondary data analysis of the Comparison of Two Screening Strategies for Gestational Diabetes trial. Participants (n = 828) self-reported the frequency of sleep disturbances (i.e., trouble falling asleep, trouble staying asleep, waking several times per night, and waking feeling tired or worn out) in mid-pregnancy. Gestational diabetes was diagnosed using either the International Associations of Diabetes and Pregnancy Study Groups or Carpenter-Coustan approach. We defined gestational diabetes subtypes based on the degree of insulin resistance and beta-cell dysfunction. We used multinomial logistic regression to examine associations of sleep disturbances with gestational diabetes status (i.e., normal, mild glycemic dysfunction, and gestational diabetes) and gestational diabetes subtypes (i.e., neither insulin resistance or beta-cell dysfunction, insulin resistance only, beta-cell dysfunction only, and insulin resistance and beta-cell dysfunction).

**Results:** A total of 665 participants (80%) had normal glycemia, 81 (10%) mild hyperglycemia, and 80 (10%) had gestational diabetes. Among participants with gestational diabetes, 62 (78%) had both insulin resistance and beta-cell dysfunction, 15 (19 %) had insulin resistance only, and 3 had beta-cell dysfunction only or neither insulin resistance nor beta-cell dysfunction. Sleep disturbance frequency was not associated with maternal glycemia or gestational diabetes subtypes.

**Conclusions:** Sleep disturbances in mid-pregnancy were not associated with maternal glycemia during mid-pregnancy. Future research should collect data on sleep disturbances at multiple time points in pregnancy and in combination with other sleep disturbances to determine whether sleep plays any role in maternal glycemic control.

## Introduction

Maternal glycemia is an important maternal health indicator during pregnancy. Hyperglycemia in pregnancy is associated with increased risk for short and long-term adverse health outcomes for the mother and child.^1^ Gestational diabetes mellitus (GDM), hyperglycemia first indicated during pregnancy, is also associated with a higher risk for pregnancy-induced hypertension, preeclampsia, and impaired glucose tolerance in the child.^2–4^ GDM can be further characterized by subtype based on the degree of insulin resistance and beta-cell dysfunction,^5, 6^ which in combination are associated with a higher risk for adverse pregnancy outcomes.^5, 6^ Hyperglycemia treatment typically includes lifestyle changes and pharmacologic therapy if needed.^7, 8^ Lifestyle changes often focus on dietary modifications and increasing physical activity.^8^ Generally, lifestyle treatment strategies do not address sleep disturbances.

Sleep disturbances, including poor sleep quality and insufficient sleep duration are common in pregnancy and may contribute to impaired maternal hyperglycemia.^9, 10^ The mechanisms linking sleep disturbances in pregnancy to maternal hyperglycemia include increased sympathetic nervous system activity, increased cortisol levels, increased inflammatory cytokine production, disruption of metabolic hormones (e.g., leptin, ghrelin) secretion, and reduced insulin sensitivity.^11–13^ For example, a meta-analysis of 15 studies found that participants with short and long sleep duration (vs. normal) had 0.23% (95% CI, 0.10–0.36) and 0.13% (95% CI, 0.02–0.25) higher hemoglobin A1C levels, respectively.^14^ Likewise, participants with poor sleep quality (vs. good) had 0.35% (95% CI, 0.12–0.58) higher hemoglobin A1C levels.

In pregnancy, epidemiological data have consistently shown strong associations between sleep quality, sleep duration, and maternal glycemia.^13, 15, 16^ However, there is little research examining associations between other sleep disturbances, such as difficulty falling asleep and staying asleep with maternal hyperglycemia.^17^ Targeting these specific sleep disturbances could potentially provide a novel approach to managing hyperglycemia in pregnancy. Therefore, this study aims to examine the associations of mid-pregnancy sleep disturbances and mid-pregnancy maternal glycemia and GDM subtypes.

## Materials and Methods

### Study Design and Population

This is a secondary data analysis of the Comparison of Two Screening Strategies for Gestational Diabetes (GDM2) trial. Trial’s design are described in detail elsewhere.^18, 19^ Briefly, the GDM2 trial was a single-center, parallel-arm, comparative effectiveness trial that aimed to compare the rates of perinatal outcomes among women randomized to receive either the International Associations of Diabetes and Pregnancy Study Groups (IADPSG) or Carpenter-Coustan GDM screening approach.^18, 19^ Participants were recruited from 10 obstetrics clinics affiliated with the University of Pittsburgh Medical Center Magee-Women’s Hospital between June 2015 and February 2019. The University of Pittsburgh’s Institutional Review Board approved the study design and procedures. Participants provided written informed consent before enrollment. An independent data safety monitoring board within the University of Pittsburgh Clinical and Translational Science Institute provided oversight. GDM2 Trial was registered at clinicaltrials.gov (NCT02309138).

Participants were recruited before routine screening for GDM between 18 to 28 weeks of gestation. Women were eligible if they were between the ages of 18-45 years and had a singleton pregnancy at screening. Exclusion criteria included: 1) preexisting type 1 or 2 diabetes mellitus, 2) diabetes diagnosed <24 weeks gestation, 3) hypertension requiring medication, 5) corticosteroid use in past 30 days, 6) anticipated preterm delivery due to maternal or fetal indications before 34 weeks gestation, 7) advanced HIV, 8) severe liver disease, or 9) gastric bypass surgery or other illness that precluded the participant from drinking the glucola solution. For this secondary analysis, we also excluded women with missing data on GDM and insomnia symptoms (n=93).

At baseline, between 24 0/7 and 28 6/7 weeks of gestation, participants in both groups received a non-fasting 50g glucola solution to drink in 5 to 10 minutes, followed by a blood draw one hour after completing the glucola solution. Participants with a blood glucose value <200 mg/dL were then randomized at a 1:1 ratio to receive a two-hour 75 g (IADPSG) or three-hour 100 g (Carpenter-Coustan) diagnostic test. Participants with 50-g OGTT values of 200 mg/dL or higher (11.1mmol/L or higher) were presumed to have gestational diabetes and excluded from the study. At visit 2, between 25 and 32 weeks of gestation, a fasting blood sample was collected for insulin, glucose, insulin resistance (HOMA-IR), and beta-cell function (HOMA-b) to evaluate baseline metabolic profiles.^18^ Participants then received either the 2-hour 75-g or 3-hour 100-g OGTT.

### Sleep Disturbances

Also at visit 2, participants were asked how often they experienced trouble falling asleep, trouble staying asleep, waking several times per night, and waking after their usual amount of sleep feeling tired or worn out in the month. Scores for each question ranged from 0 (not at all) to 5 (22 to 31 days). The total frequency of sleep disturbances was defined as the sum of each question (possible range of 0 to 20), with higher scores indicating more frequent sleep disturbances.

### Maternal Glycemia and GDM subtypes

A central clinical laboratory at UPMC performed all the glucose tests and communicated the results with the study team to make GDM diagnosis based on either the IADPSG or Carpenter-Coustan approach. GDM was diagnosed in the IADPSG group if the fasting 75-g OGTT had at least one abnormal value (fasting 92 mg/dL or higher [5.11 mmol/L], 1-hour 180 or higher [9.99], 2-hour 153 or higher [8.49]); the 50-g OGTT was ignored. GDM was diagnosed in the Carpenter-Coustan group if the 50-g OGTT value was 130 mg/dL or higher (7.215 mmol/L or higher) and the fasting 100-g OGTT had two or more abnormal values (fasting 95 mg/dL or higher [5.27 mmol/L], 1-hour 180 or higher [9.99]; 2-hour 155 or higher [8.60]; 3-hour 140 or higher [7.77]). We defined “mild” glycemic dysfunction in the Carpenter-Coustan group as having only one abnormal fasting glucose test (i.e., 50 gm <130 mg/dL OR 50 gm >130 mg/d AND <1 abnormal value from 100 gm test).

We defined glycemic subtypes based on the degree of insulin resistance and beta-cell dysfunction. We calculated HOMA-IR (i.e., fasting insulin (μIU/ml) x fasting glucose (mg/dl)/405) and HOMA-B (i.e., 360 × fasting insulin (μIU/ml)/fasting glucose (mg/dl) − 63) scores to estimate insulin resistance and beta-cell dysfunction, respectively. We used the distribution of insulin and beta-cell function values among women with normal glycemia and beta-cell dysfunction to define our insulin resistance and beta-cell dysfunction groups.

Specifically, insulin resistance and beta-cell dysfunction were defined as HOMA-IR and HOMA-B values <25^th^ percentile in the sample distribution, respectively. Next, we explored the presence of the following glycemic subtypes; 1) normal glycemia, 2) mild glycemic dysfunction, 2) GDM with insulin resistance, 4) GDM with beta-cell dysfunction, and 5) GDM with insulin resistance and beta-cell dysfunction, and 6) GDM with neither insulin resistance nor beta-cell dysfunction. Few people (n=1) were classified as GDM with beta-cell dysfunction alone or GDM without insulin resistance or beta-cell dysfunction (n=1). Thus, our final analytic sample included participants with normal, mild insulin resistance, and insulin resistance + beta-cell dysfunction

### Covariates

Covariate data were collected at baseline and included demographic, behavioral, psychosocial, and clinical risk factors for maternal hyperglycemia. Demographic characteristics included age and race/ethnicity. Participants self-reported their race as American Indian/Alaska Native, Asian, Native Hawaiian or other Pacific Islander, Black, White, or more than one race. Participants were also asked if they were Hispanic or Latinx. We grouped participants according to the US census as Asian, White, Black, and “all other races” because of small sample sizes.

Participants also self-reported their employment status (part-time, full-time, receiving disability, unemployed), education, marital status, income, the number of adults in the household, and current smoking and alcohol intake status.

Clinical characteristics included pre-pregnancy BMI, gestational weight gain, and current health status. Appropriate, excess, and inadequate gestational weight gain was defined based on Institute of Medicine guidelines. Adjusted maternal weight gain was defined as total weight gain minus infant birthweight to account for the contribution of fetal weight to weight gain.

Psychosocial factors included depressive symptoms and perceived stress. The Edinburgh Postnatal Depression Scale (EPDS) was used to measure depression symptoms.^20^ This 10-item self-reported scale identifies symptoms of depression such as feeling sad or miserable, the inability to cope, and sleep disruption in the past week. The scores range from 0 to 30, with higher scores indicating more depressive symptoms. An EPDS score of 13 or higher promoted us to provide supportive resources for depression. The scale has acceptable sensitivity (86%) and specificity (78%) for predicting depression. The Perceived Stress Scale (PSS)^21^ is a 14-item scale used to assess how often participants experienced stress in the past month. Scores range from 0 to 56, with higher scores indicating more perceived stress. In a general population, the test-retest correlations ranged between 0.55 and 0.85.^21^

### Statistical Analysis

Descriptive statistics such as means, standard deviations, and proportions were calculated for participant demographics and baseline clinical and psychosocial characteristics. Our cohort was grouped into tertiles based on the possible values of the sleep disturbance frequency (i.e., 0- 6, 7-13, and 14-20). Chi-Square test, Analysis of Variance, or nonparametric equivalents (Fisher’s Exact or Kruskal-Wallis tests) were used to compare baseline characteristics across insomnia frequency groups.

For the primary analysis, we used multinomial logistic regression to examine associations of sleep disturbances (both as a continuous value and tertile group) with GDM status (i.e., normal, mild glycemic dysfunction, GDM). Odds ratios (ORs) and 95% confidence intervals (CI) were constructed based on the following comparisons: mild vs. normal and GDM vs. normal. For each model, the lowest sleep disturbance frequency group (Not at all) or the lowest tertile of overall sleep disturbance was treated as the reference group. Additionally, we ran adjusted models, including covariates related to the sleep disturbance frequency group from Table 1. A similar analysis strategy was used to model the association between sleep disturbances and maternal glycemia subtype (normal, mild, insulin resistance, and insulin resistance + beta-cell dysfunction).

**Table 1:**
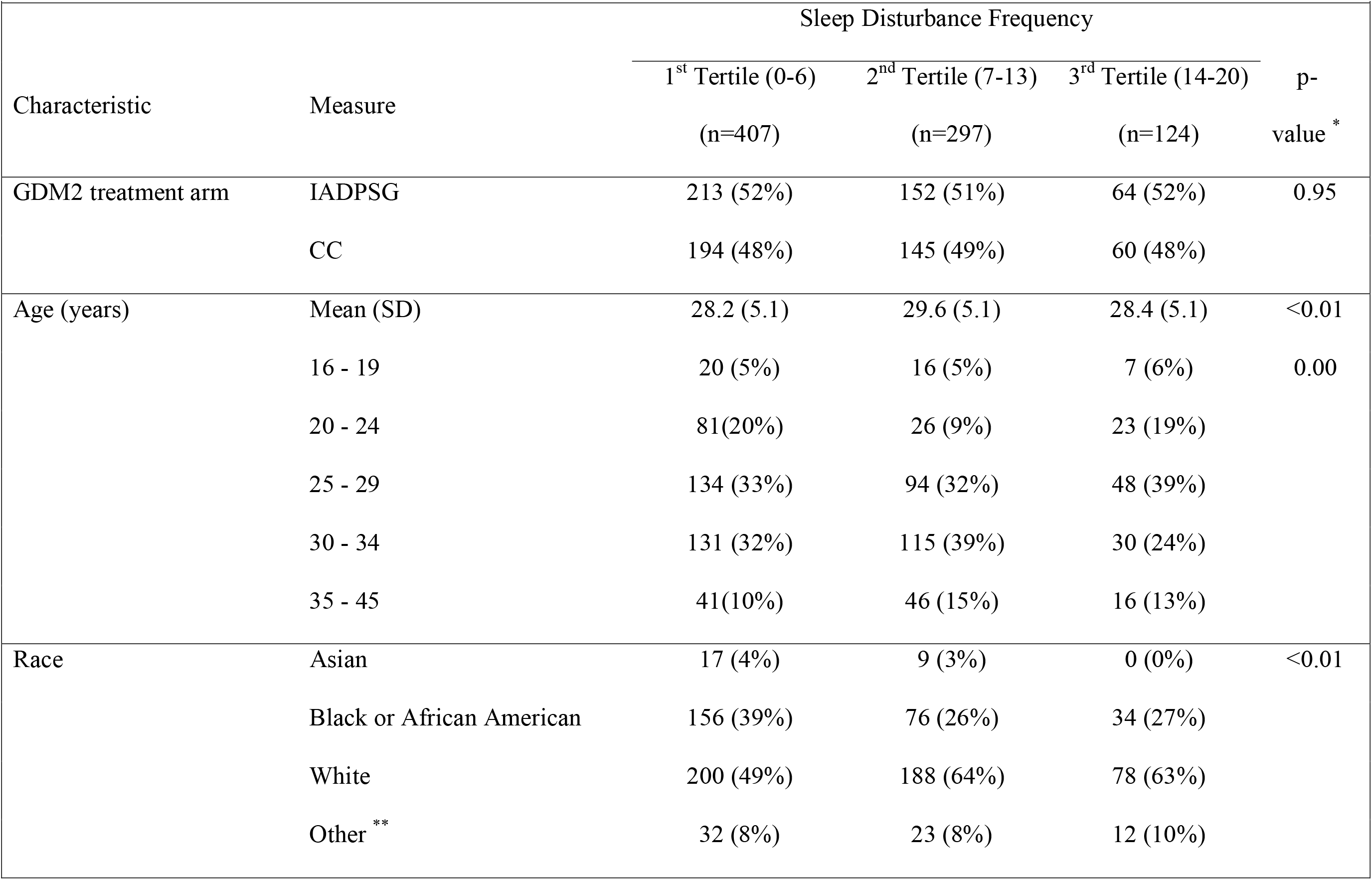
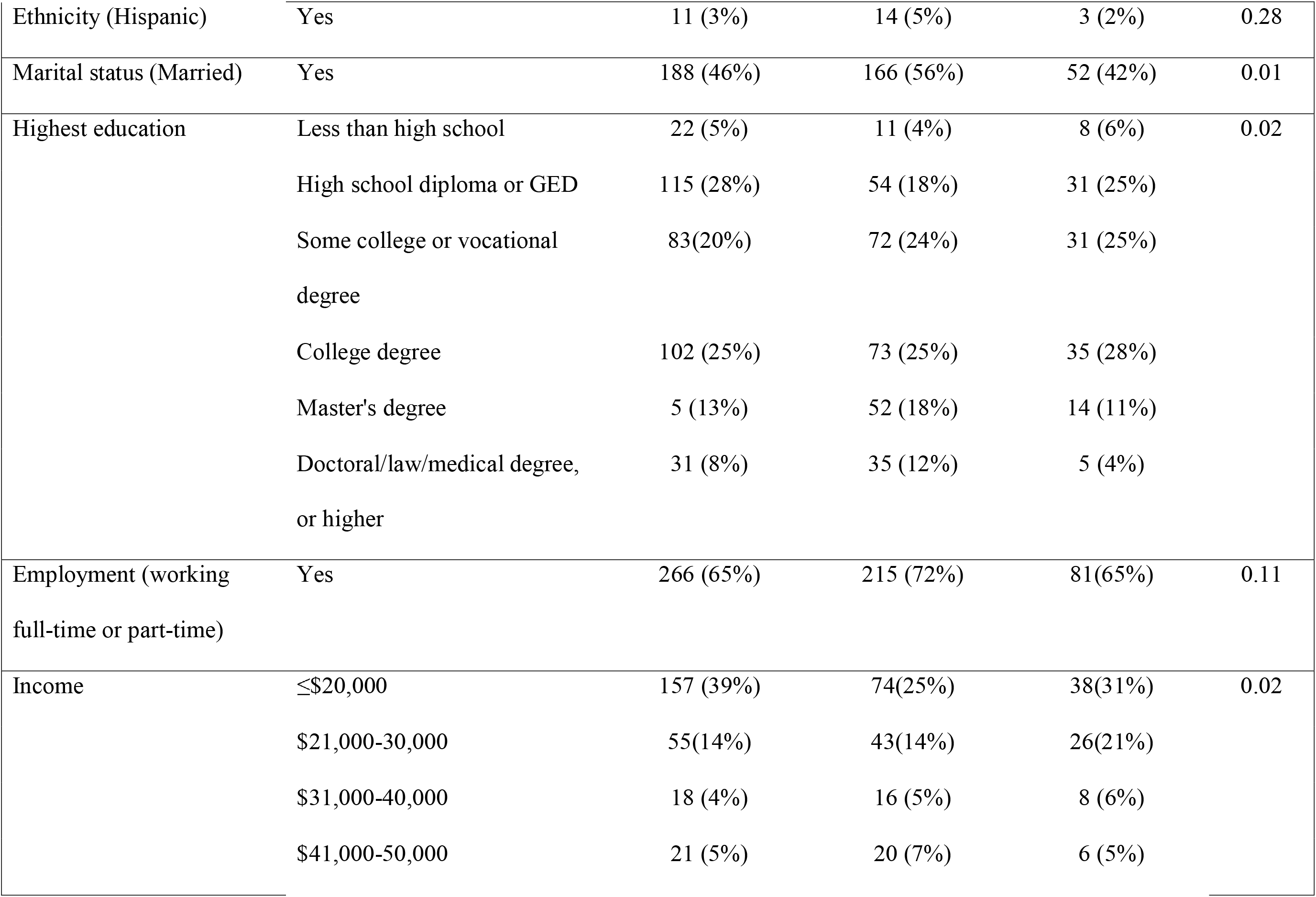
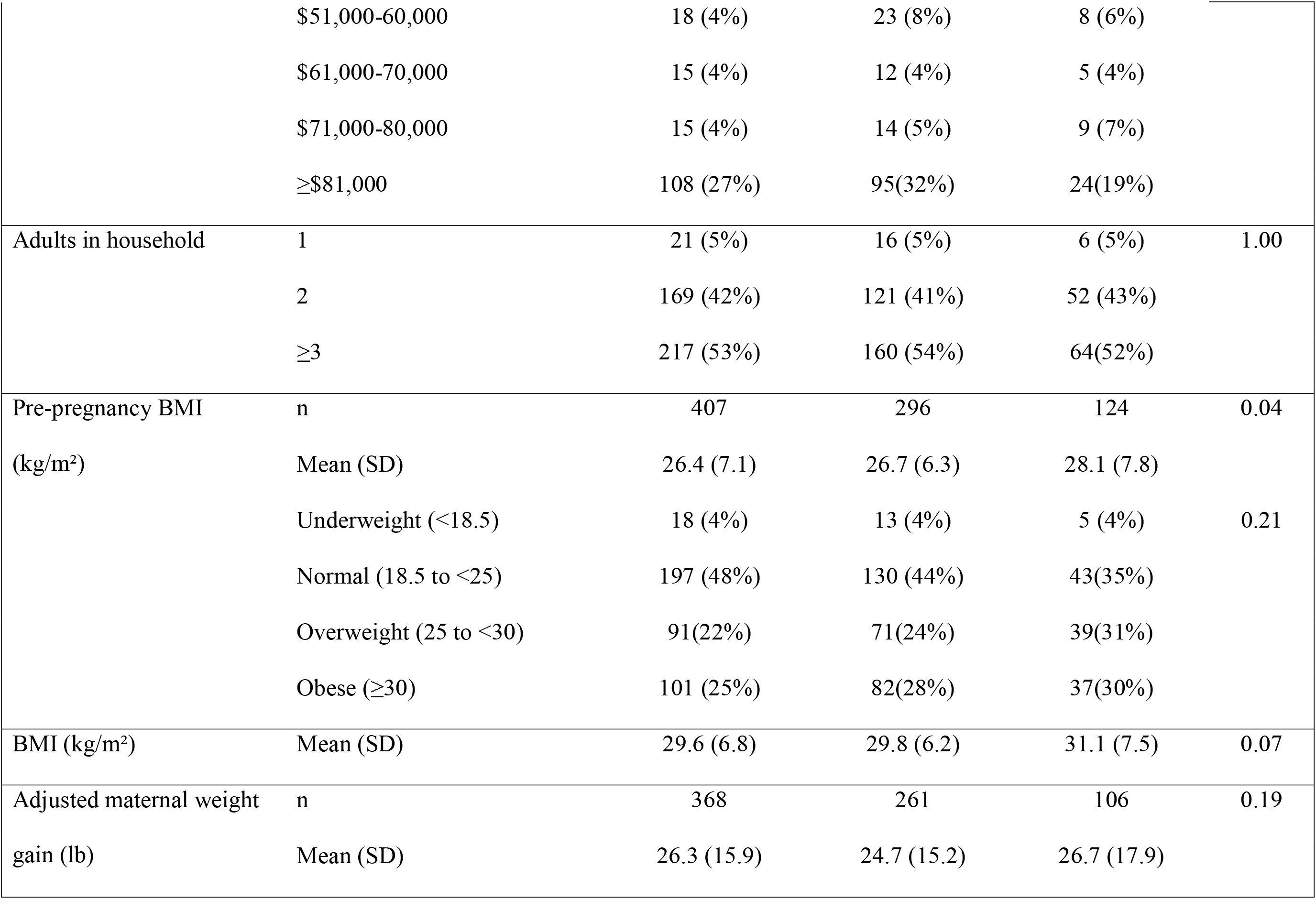
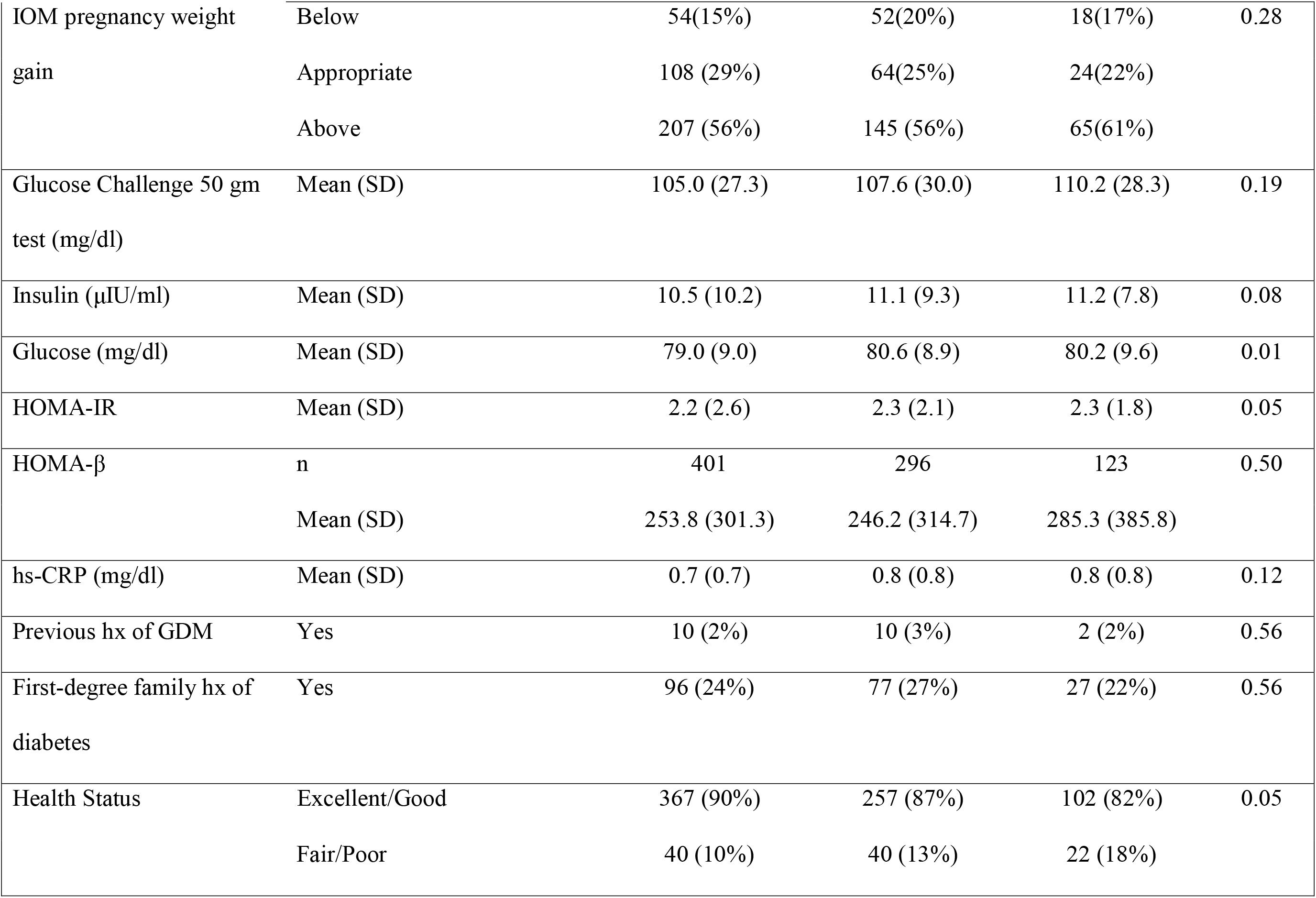
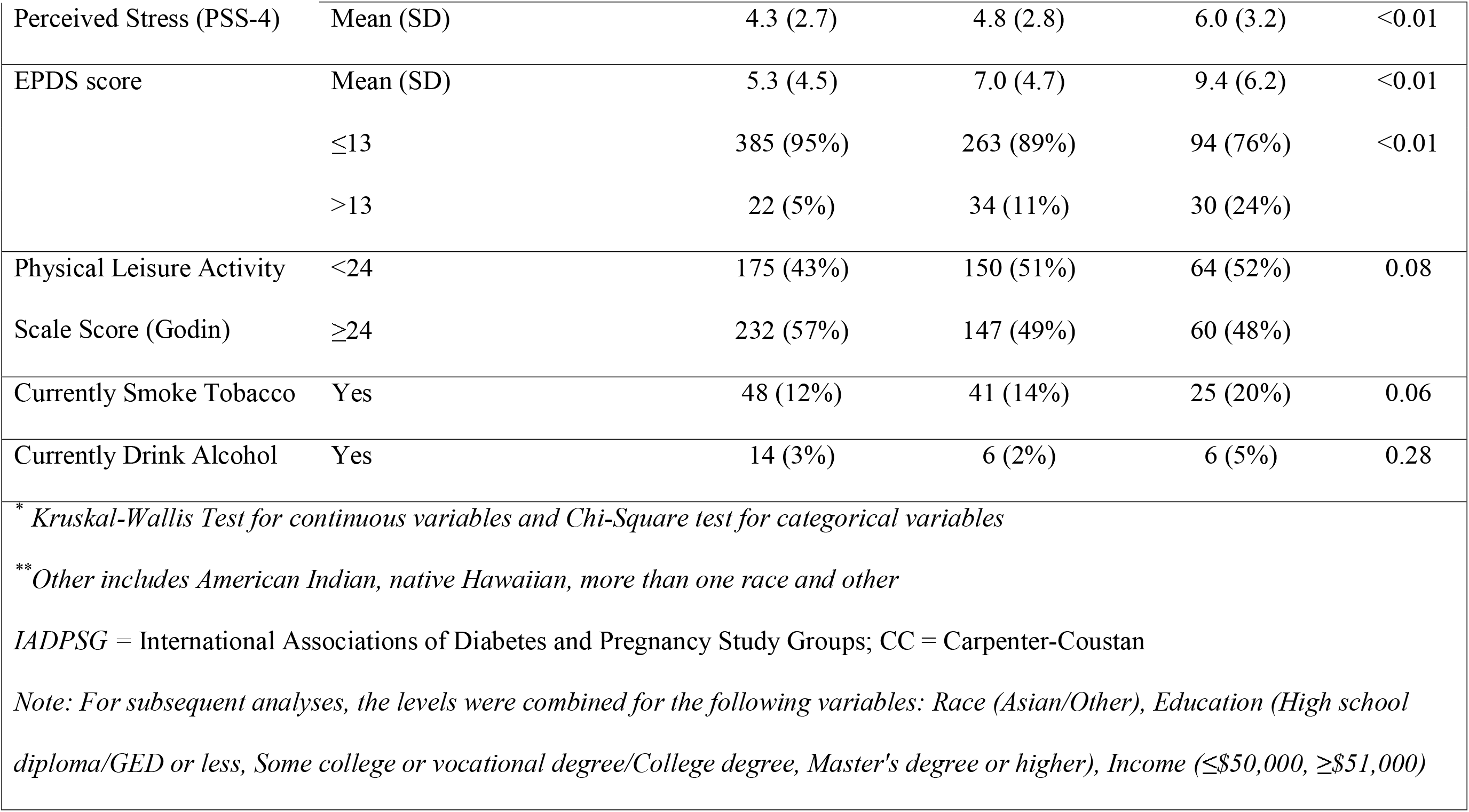
Participant characteristics by sleep disturbance frequency

| Table 1: Participant characteristics by sleep disturbance frequency |  |  |  |  |  |
| --- | --- | --- | --- | --- | --- |
| Characteristic | Measure | Sleep Disturbance Frequency |  |  | p-value * |
|  |  | 1<sup>st</sup> Tertile (0-6)<br>(n=407) | 2<sup>nd</sup> Tertile (7-13)<br>(n=297) | 3<sup>rd</sup> Tertile (14-20)<br>(n=124) |  |
| GDM2 treatment arm | IADPSG | 213 (52%) | 152 (51%) | 64 (52%) | 0.95 |
|  | CC | 194 (48%) | 145 (49%) | 60 (48%) |  |
| Age (years) | Mean (SD) | 28.2 (5.1) | 29.6 (5.1) | 28.4 (5.1) | <0.01 |
|  | 16 - 19 | 20 (5%) | 16 (5%) | 7 (6%) | 0.00 |
|  | 20 - 24 | 81(20%) | 26 (9%) | 23 (19%) |  |
|  | 25 - 29 | 134 (33%) | 94 (32%) | 48 (39%) |  |
|  | 30 - 34 | 131 (32%) | 115 (39%) | 30 (24%) |  |
|  | 35 - 45 | 41(10%) | 46 (15%) | 16 (13%) |  |
| Race | Asian | 17 (4%) | 9 (3%) | 0 (0%) | <0.01 |
|  | Black or African American | 156 (39%) | 76 (26%) | 34 (27%) |  |
|  | White | 200 (49%) | 188 (64%) | 78 (63%) |  |
|  | Other ** | 32 (8%) | 23 (8%) | 12 (10%) |  |
| Ethnicity (Hispanic) | Yes | 11 (3%) | 14 (5%) | 3 (2%) | 0.28 |
| Marital status (Married) | Yes | 188 (46%) | 166 (56%) | 52 (42%) | 0.01 |
| Highest education | Less than high school | 22 (5%) | 11 (4%) | 8 (6%) | 0.02 |
|  | High school diploma or GED | 115 (28%) | 54 (18%) | 31 (25%) |  |
|  | Some college or vocational degree | 83(20%) | 72 (24%) | 31 (25%) |  |
|  | College degree | 102 (25%) | 73 (25%) | 35 (28%) |  |
|  | Master's degree | 5 (13%) | 52 (18%) | 14 (11%) |  |
|  | Doctoral/law/medical degree, or higher | 31 (8%) | 35 (12%) | 5 (4%) |  |
| Employment (working full-time or part-time) | Yes | 266 (65%) | 215 (72%) | 81(65%) | 0.11 |
| Income | ≤\$20,000 | 157 (39%) | 74(25%) | 38(31%) | 0.02 |
| | \$21,000-30,000 | 55(14%) | 43(14%) | 26(21%) | |
| | \$31,000-40,000 | 18 (4%) | 16 (5%) | 8 (6%) | |
| | \$41,000-50,000 | 21 (5%) | 20 (7%) | 6 (5%) | |
| | \$51,000-60,000 | 18 (4%) | 23 (8%) | 8 (6%) | |
| | \$61,000-70,000 | 15 (4%) | 12 (4%) | 5 (4%) | |
| | \$71,000-80,000 | 15 (4%) | 14 (5%) | 9 (7%) | |
| | ≥\$81,000 | 108 (27%) | 95(32%) | 24(19%) | |
| Adults in household | 1 | 21 (5%) | 16 (5%) | 6 (5%) | 1.00 |
|  | 2 | 169 (42%) | 121 (41%) | 52 (43%) |  |
|  | ≥3 | 217 (53%) | 160 (54%) | 64(52%) |  |
| Pre-pregnancy BMI | n | 407 | 296 | 124 | 0.04 |
| (kg/m<sup>2</sup>) | Mean (SD) | 26.4 (7.1) | 26.7 (6.3) | 28.1 (7.8) |  |
|  | Underweight (<18.5) | 18 (4%) | 13 (4%) | 5 (4%) | 0.21 |
|  | Normal (18.5 to <25) | 197 (48%) | 130 (44%) | 43(35%) |  |
|  | Overweight (25 to <30) | 91(22%) | 71(24%) | 39(31%) |  |
|  | Obese (≥30) | 101 (25%) | 82(28%) | 37(30%) |  |
| BMI (kg/m<sup>2</sup>) | Mean (SD) | 29.6 (6.8) | 29.8 (6.2) | 31.1 (7.5) | 0.07 |
| Adjusted maternal weight | n | 368 | 261 | 106 | 0.19 |
| gain (lb) | Mean (SD) | 26.3 (15.9) | 24.7 (15.2) | 26.7 (17.9) |  |
| IOM pregnancy weight gain | Below | 54(15%) | 52(20%) | 18(17%) | 0.28 |
|  | Appropriate | 108 (29%) | 64(25%) | 24(22%) |  |
|  | Above | 207 (56%) | 145 (56%) | 65(61%) |  |
| Glucose Challenge 50 gm test (mg/dl) | Mean (SD) | 105.0 (27.3) | 107.6 (30.0) | 110.2 (28.3) | 0.19 |
| Insulin (μIU/ml) | Mean (SD) | 10.5 (10.2) | 11.1 (9.3) | 11.2 (7.8) | 0.08 |
| Glucose (mg/dl) | Mean (SD) | 79.0 (9.0) | 80.6 (8.9) | 80.2 (9.6) | 0.01 |
| HOMA-IR | Mean (SD) | 2.2 (2.6) | 2.3 (2.1) | 2.3 (1.8) | 0.05 |
| HOMA-β | n | 401 | 296 | 123 | 0.50 |
|  | Mean (SD) | 253.8 (301.3) | 246.2 (314.7) | 285.3 (385.8) |  |
| hs-CRP (mg/dl) | Mean (SD) | 0.7 (0.7) | 0.8 (0.8) | 0.8 (0.8) | 0.12 |
| Previous hx of GDM | Yes | 10 (2%) | 10 (3%) | 2 (2%) | 0.56 |
| First-degree family hx of diabetes | Yes | 96 (24%) | 77 (27%) | 27 (22%) | 0.56 |
| Health Status | Excellent/Good | 367 (90%) | 257 (87%) | 102 (82%) | 0.05 |
|  | Fair/Poor | 40 (10%) | 40 (13%) | 22 (18%) |  |
| Perceived Stress (PSS-4) | Mean (SD) | 4.3 (2.7) | 4.8 (2.8) | 6.0 (3.2) | <0.01 |
| EPDS score | Mean (SD) | 5.3 (4.5) | 7.0 (4.7) | 9.4 (6.2) | <0.01 |
|  | ≤13 | 385 (95%) | 263 (89%) | 94 (76%) | <0.01 |
|  | >13 | 22 (5%) | 34 (11%) | 30 (24%) |  |
| Physical Leisure Activity | <24 | 175 (43%) | 150 (51%) | 64 (52%) | 0.08 |
| Scale Score (Godin) | ≥24 | 232 (57%) | 147 (49%) | 60 (48%) |  |
| Currently Smoke Tobacco | Yes | 48 (12%) | 41 (14%) | 25 (20%) | 0.06 |
| Currently Drink Alcohol | Yes | 14 (3%) | 6 (2%) | 6 (5%) | 0.28 |
| * *Kruskal-Wallis Test for continuous variables and Chi-Square test for categorical variables*<br>** *Other includes American Indian, native Hawaiian, more than one race and other*<br>IADPSG = International Associations of Diabetes and Pregnancy Study Groups; CC = Carpenter-Coustan<br>*Note: For subsequent analyses, the levels were combined for the following variables: Race (Asian/Other), Education (High school diploma/GED or less, Some college or vocational degree/College degree, Master's degree or higher), Income (≤\$50,000, ≥\$51,000)* | | | | | |

Linear regression models were constructed to assess the relationship between sleep disturbance frequency and log-transformed fasting glucose. We report beta coefficients expressed as “% change” along with 95% CIs.

SAS version 9.4 (SAS Institute Inc) was used for all statistical analyses. We used an alpha of <0.05 to define statistical significance.

## Results

The final analysis sample included 828 participants with data on sleep disturbances in mid-pregnancy and maternal glycemia in mid-pregnancy. Table 1 describes baseline characteristics by tertiles of sleep disturbance frequency. A higher percentage of participants in the highest tertile (vs. the lowest tertile) of sleep disturbances were White, single or not married, had a higher pre-pregnancy BMI, reported fair or poor overall health, had higher levels of perceived stress and had a higher frequency of depressive symptoms. Women in the highest sleep disturbance tertile (vs. the lowest tertile) were also less physically active. A higher percentage of women in the highest sleep disturbance tertile reported currently smoking; however, these differences were not statistically significant.

A total of 665 (80%) participants had normal glycemia, 81 (10%) had mild glycemic dysfunction, and 80 (10%) had GDM, respectively. There were no statistically significant differences in the frequency of sleep disturbances in participants with mild glycemia dysfunction or GDM compared to participants with normal glycemic function in unadjusted or adjusted models (Table 2). Also, there were no statistically significant associations between sleep disturbance frequency and fasting glucose, HOMA-IR, or HOMA-B, except for reported trouble falling asleep (Table 3). Specifically, having trouble falling asleep >14 days in the past month (vs. < 14 days) was positively associated with HOMA-B. The association between trouble falling asleep and HOMA-B was attenuated and no longer statistically significant after covariate adjustment.

**Table 2:**
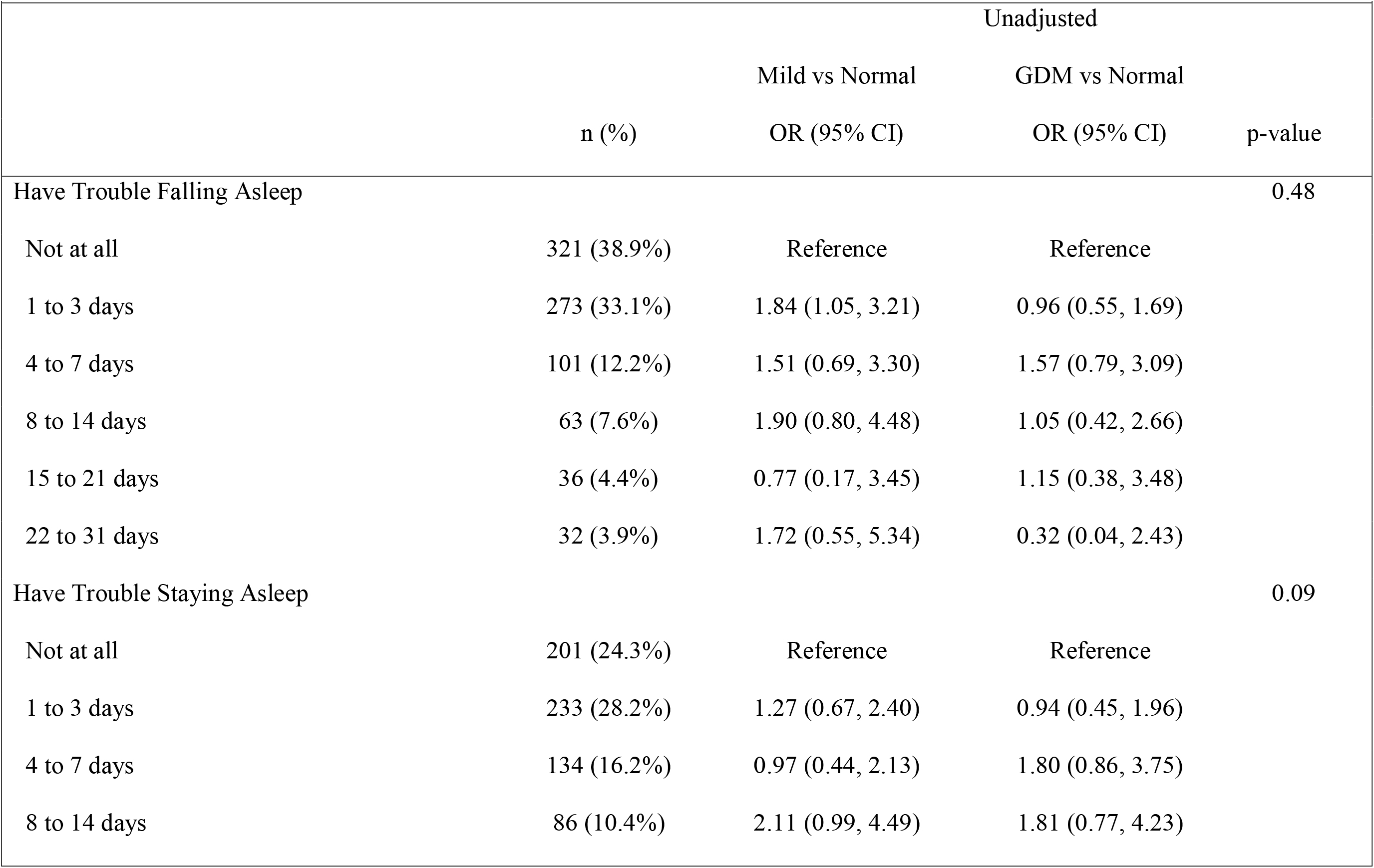
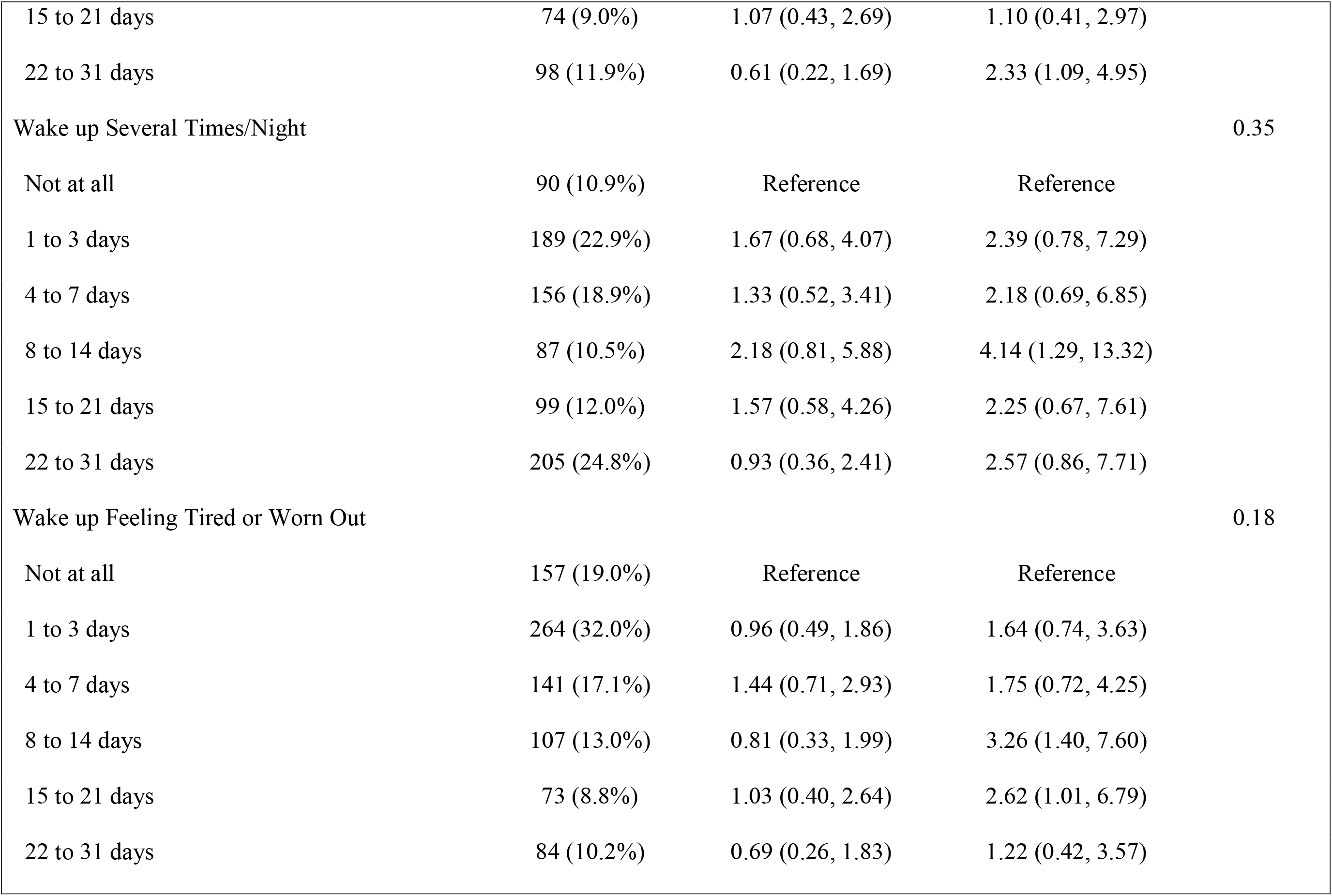
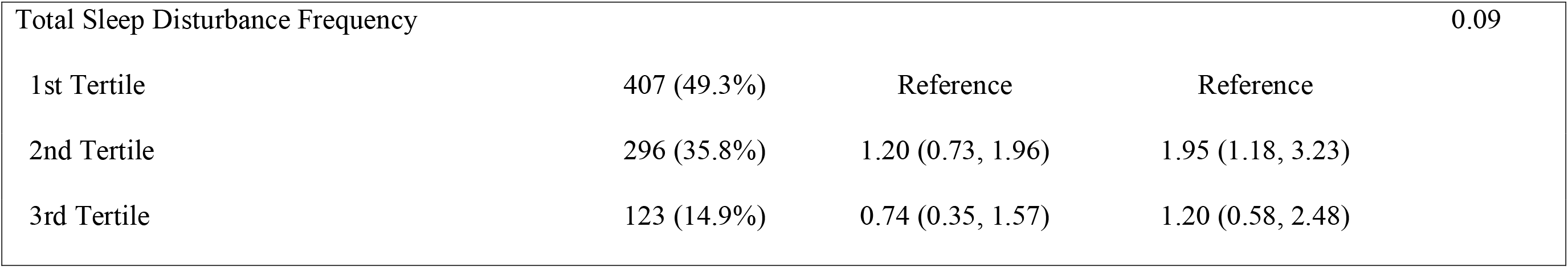
Association between maternal sleep disturbance frequency and GDM Status

|  |  |  |  |  |
| --- | --- | --- | --- | --- |
|  |  | Mild vs Normal | Unadjusted |  |
|  | n (%) | OR (95% CI) | GDM vs Normal<br>OR (95% CI) | p-value |
| Have Trouble Falling Asleep |  |  |  | 0.48 |
| Not at all | 321 (38.9%) | Reference | Reference |  |
| 1 to 3 days | 273 (33.1%) | 1.84 (1.05, 3.21) | 0.96 (0.55, 1.69) |  |
| 4 to 7 days | 101 (12.2%) | 1.51 (0.69, 3.30) | 1.57 (0.79, 3.09) |  |
| 8 to 14 days | 63 (7.6%) | 1.90 (0.80, 4.48) | 1.05 (0.42, 2.66) |  |
| 15 to 21 days | 36 (4.4%) | 0.77 (0.17, 3.45) | 1.15 (0.38, 3.48) |  |
| 22 to 31 days | 32 (3.9%) | 1.72 (0.55, 5.34) | 0.32 (0.04, 2.43) |  |
| Have Trouble Staying Asleep |  |  |  | 0.09 |
| Not at all | 201 (24.3%) | Reference | Reference |  |
| 1 to 3 days | 233 (28.2%) | 1.27 (0.67, 2.40) | 0.94 (0.45, 1.96) |  |
| 4 to 7 days | 134 (16.2%) | 0.97 (0.44, 2.13) | 1.80 (0.86, 3.75) |  |
| 8 to 14 days | 86 (10.4%) | 2.11 (0.99, 4.49) | 1.81 (0.77, 4.23) |  |
| 15 to 21 days | 74 (9.0%) | 1.07 (0.43, 2.69) | 1.10 (0.41, 2.97) |  |
| 22 to 31 days | 98 (11.9%) | 0.61 (0.22, 1.69) | 2.33 (1.09, 4.95) |  |
| Wake up Several Times/Night |  |  |  | 0.35 |
| Not at all | 90 (10.9%) | Reference | Reference |  |
| 1 to 3 days | 189 (22.9%) | 1.67 (0.68, 4.07) | 2.39 (0.78, 7.29) |  |
| 4 to 7 days | 156 (18.9%) | 1.33 (0.52, 3.41) | 2.18 (0.69, 6.85) |  |
| 8 to 14 days | 87 (10.5%) | 2.18 (0.81, 5.88) | 4.14 (1.29, 13.32) |  |
| 15 to 21 days | 99 (12.0%) | 1.57 (0.58, 4.26) | 2.25 (0.67, 7.61) |  |
| 22 to 31 days | 205 (24.8%) | 0.93 (0.36, 2.41) | 2.57 (0.86, 7.71) |  |
| Wake up Feeling Tired or Worn Out |  |  |  | 0.18 |
| Not at all | 157 (19.0%) | Reference | Reference |  |
| 1 to 3 days | 264 (32.0%) | 0.96 (0.49, 1.86) | 1.64 (0.74, 3.63) |  |
| 4 to 7 days | 141 (17.1%) | 1.44 (0.71, 2.93) | 1.75 (0.72, 4.25) |  |
| 8 to 14 days | 107 (13.0%) | 0.81 (0.33, 1.99) | 3.26 (1.40, 7.60) |  |
| 15 to 21 days | 73 (8.8%) | 1.03 (0.40, 2.64) | 2.62 (1.01, 6.79) |  |
| 22 to 31 days | 84 (10.2%) | 0.69 (0.26, 1.83) | 1.22 (0.42, 3.57) |  |
| Total Sleep Disturbance Frequency |  |  |  | 0.09 |
| 1st Tertile | 407 (49.3%) | Reference | Reference |  |
| 2nd Tertile | 296 (35.8%) | 1.20 (0.73, 1.96) | 1.95 (1.18, 3.23) |  |
| 3rd Tertile | 123 (14.9%) | 0.74 (0.35, 1.57) | 1.20 (0.58, 2.48) |  |

**Table 3:**
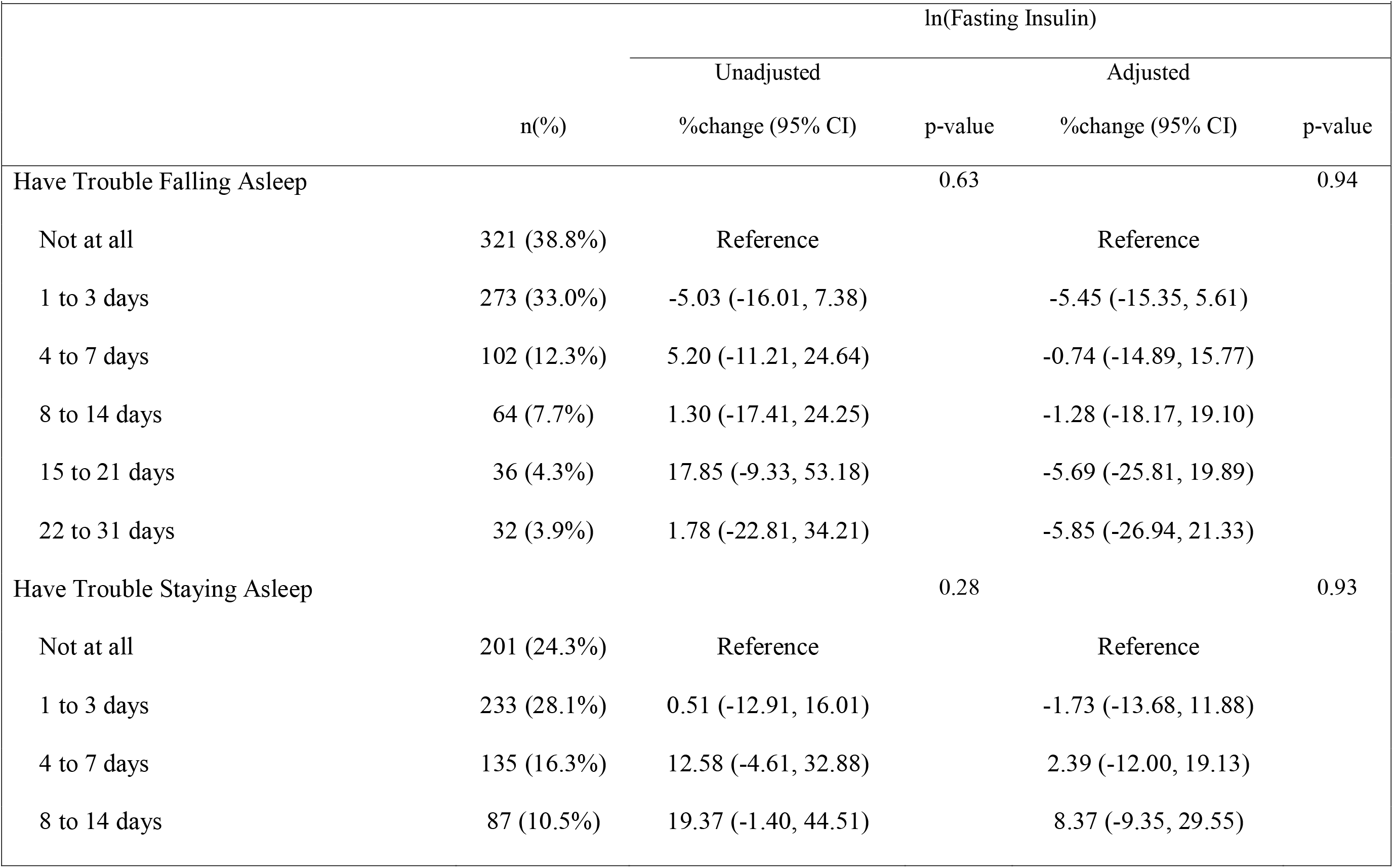
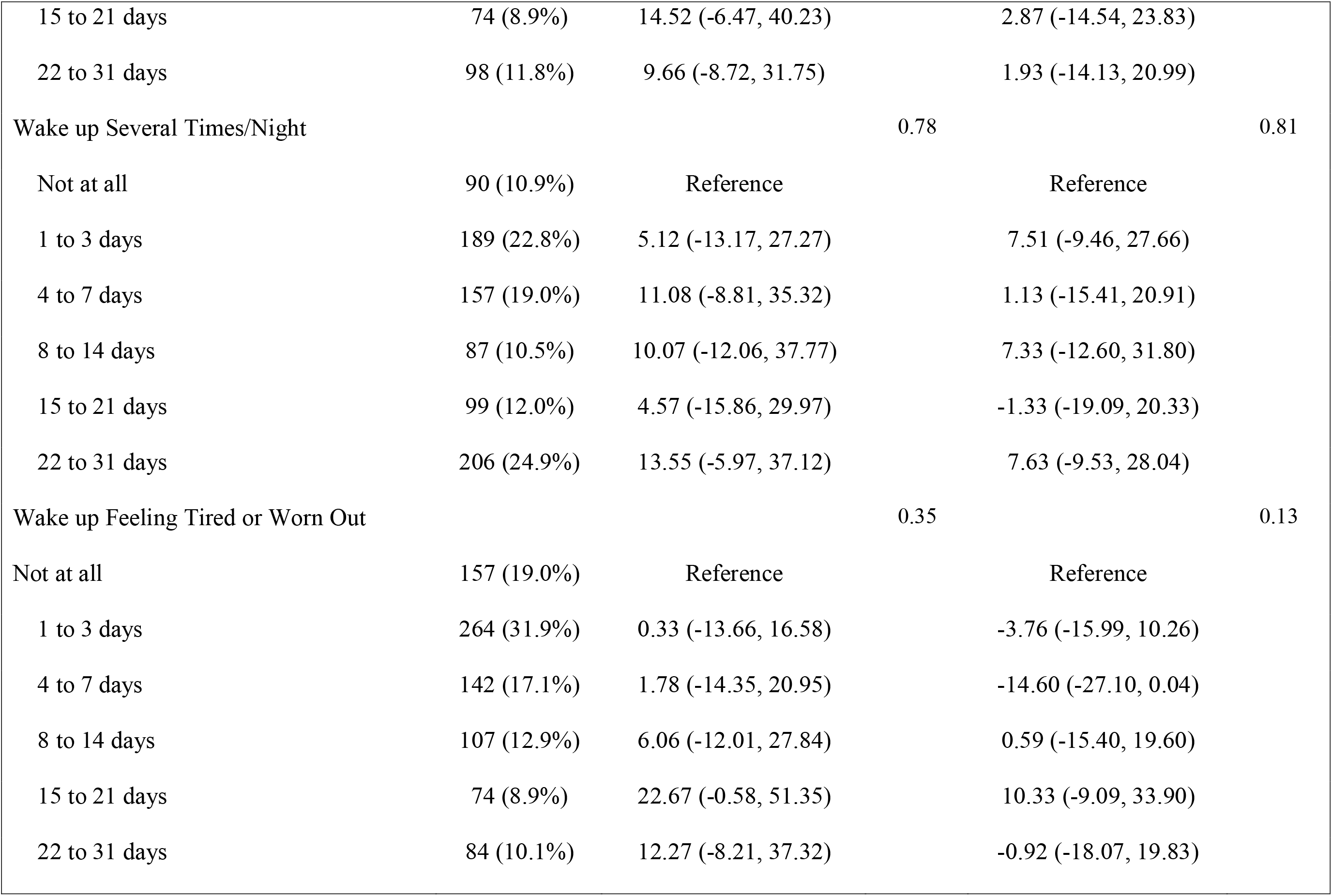
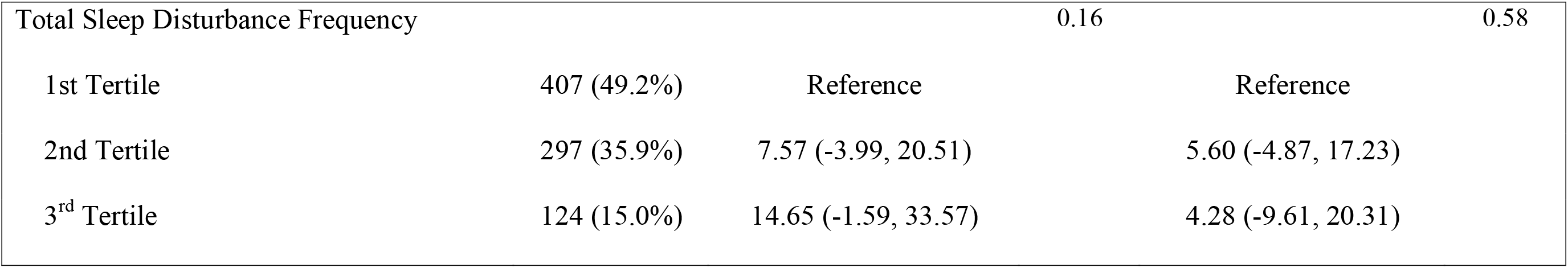
Association between maternal sleep disturbances and fasting insulin

Next, we examined associations between the frequency of sleep disturbances and GDM subtypes (Table 4). There were no statistically significant associations between sleep disturbance frequency and GDM subtype, except for having trouble staying asleep. This association was attenuated and no longer statistically significant after covariate adjustment.

**Table 4:**
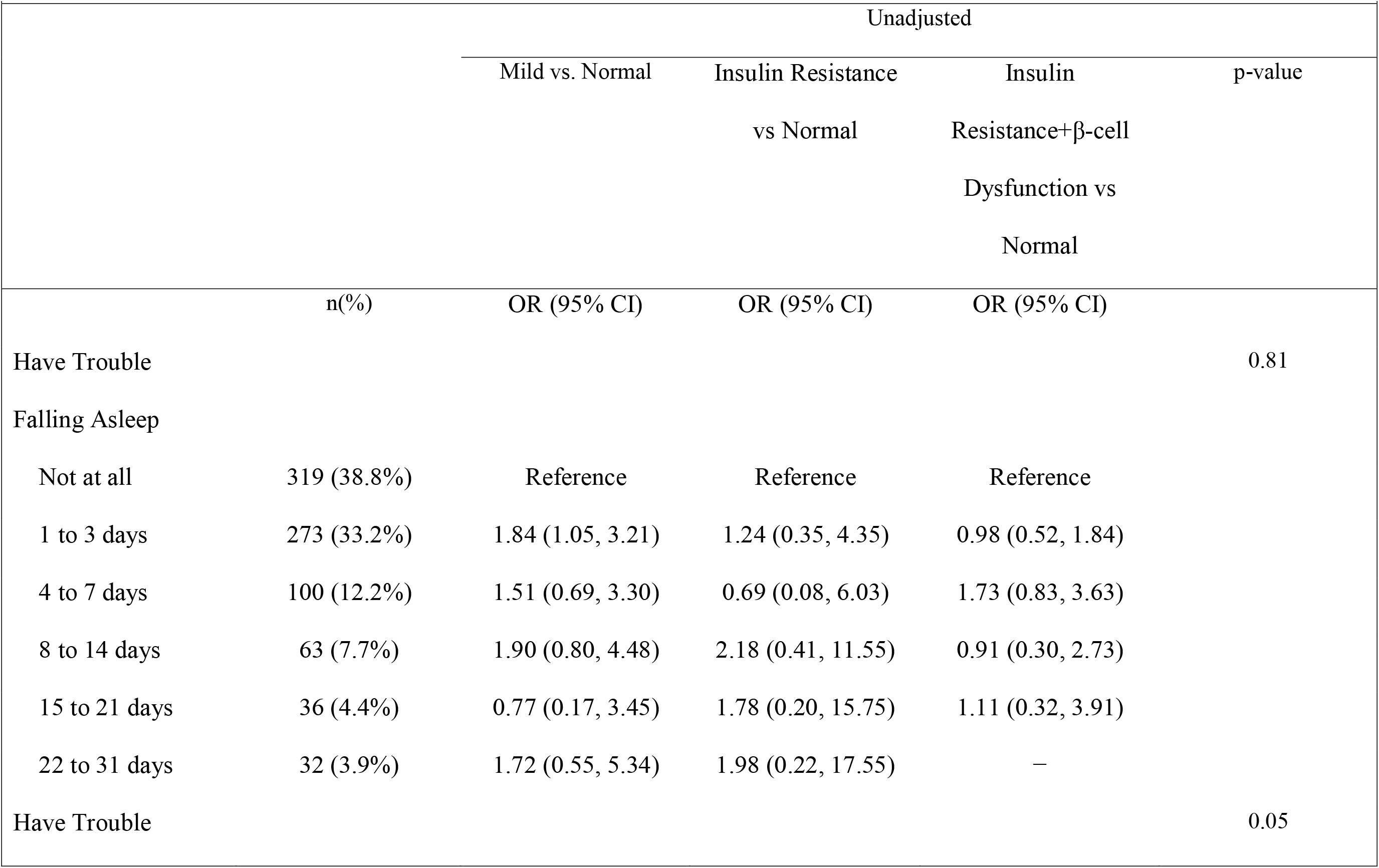
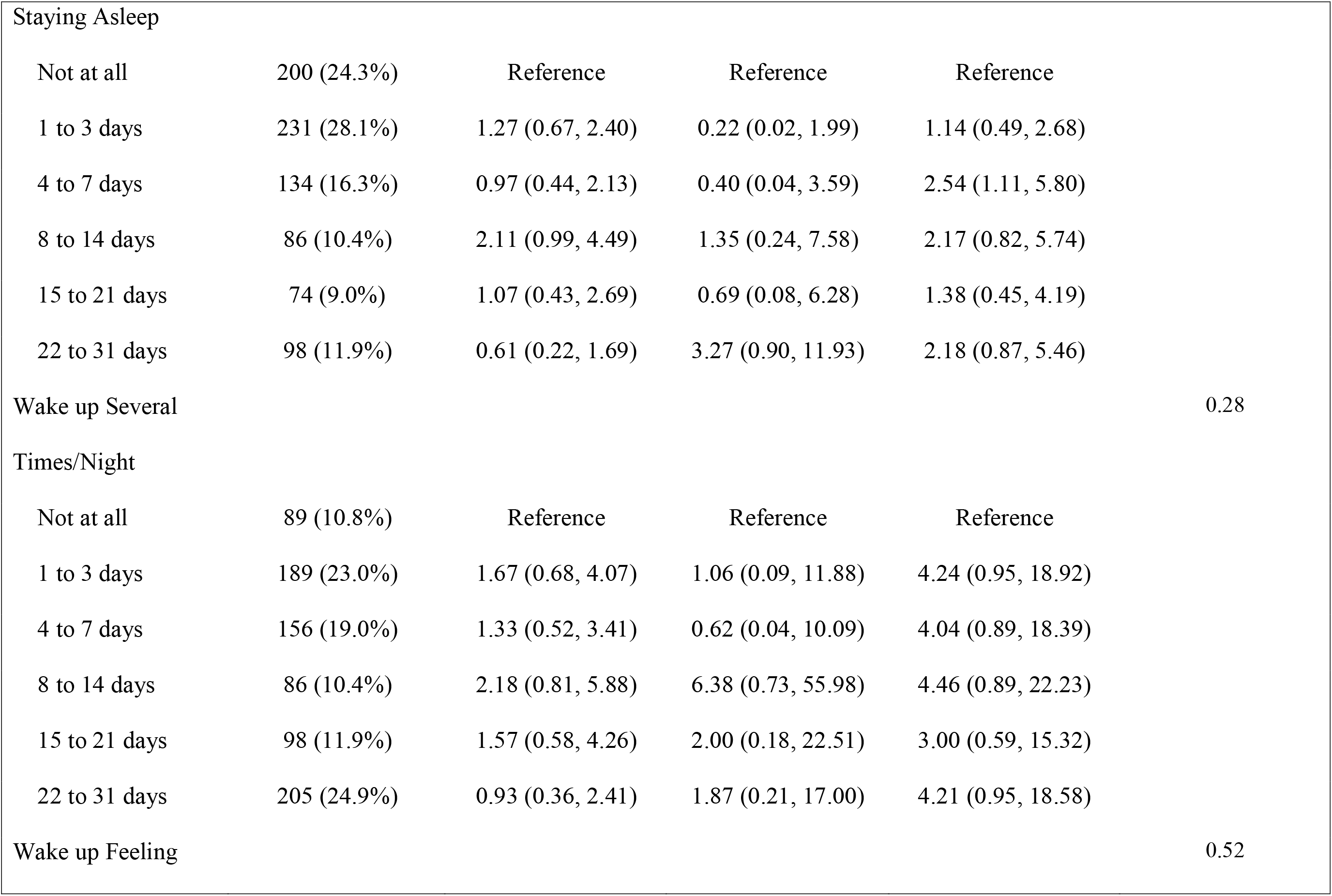
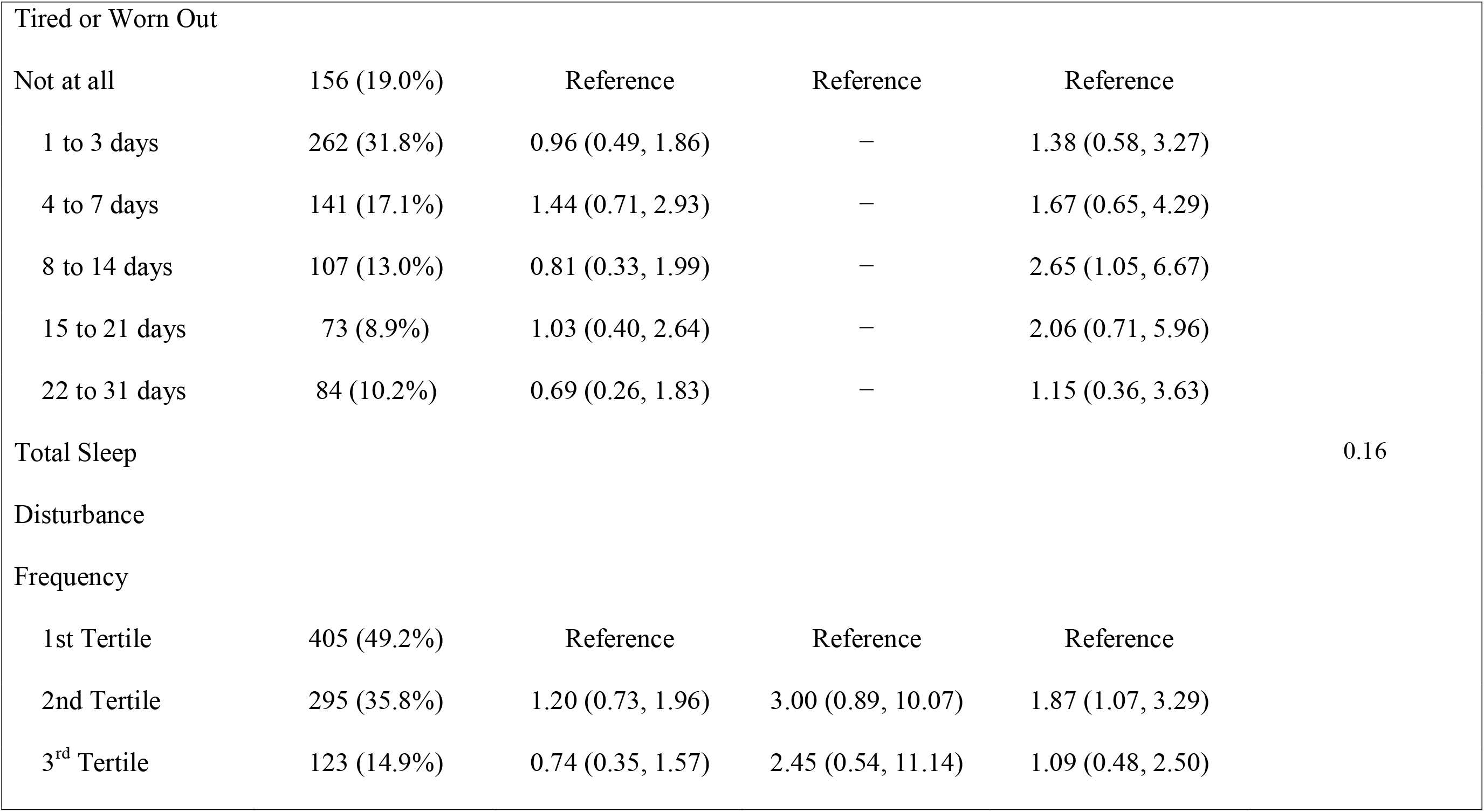
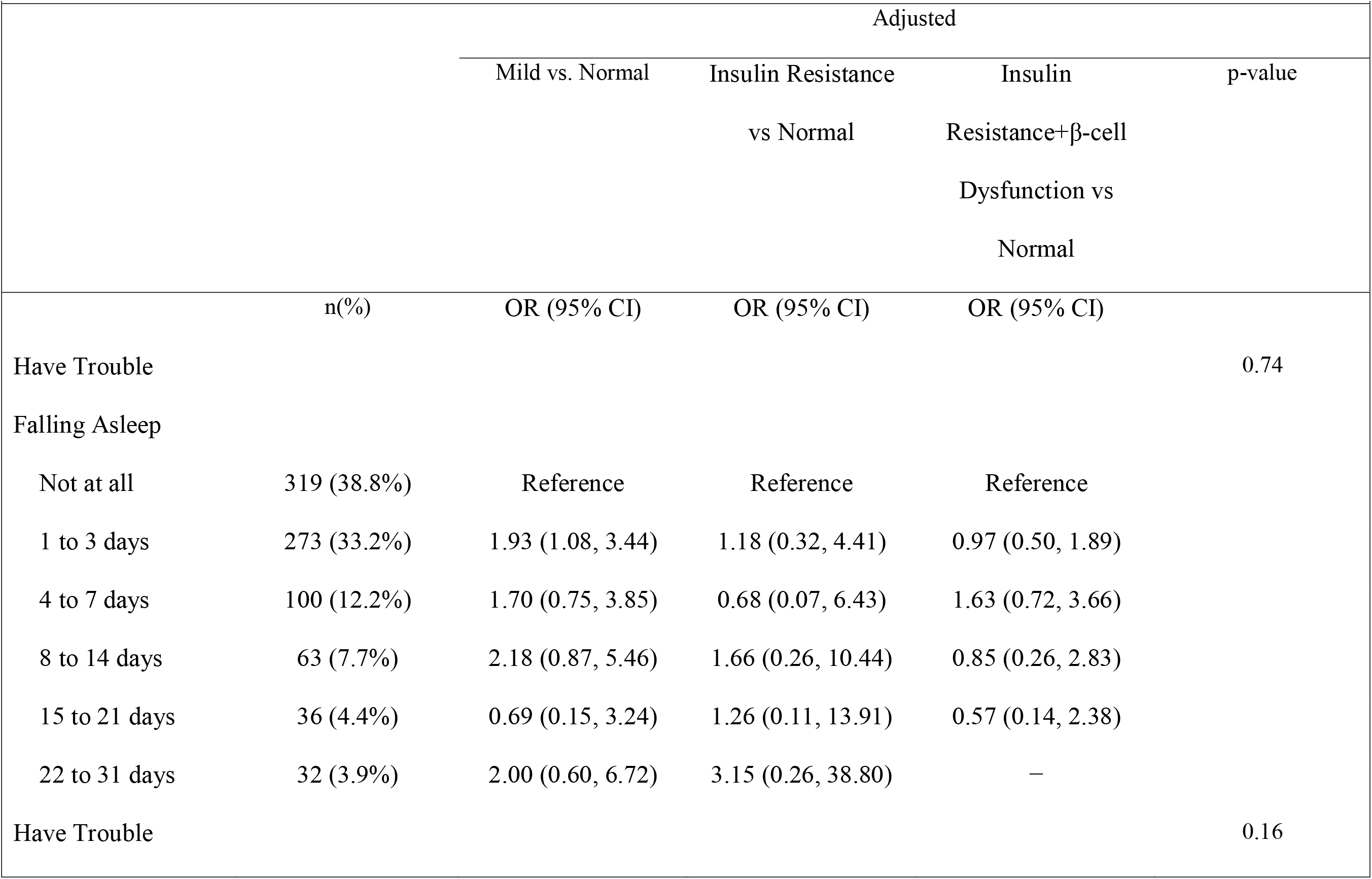
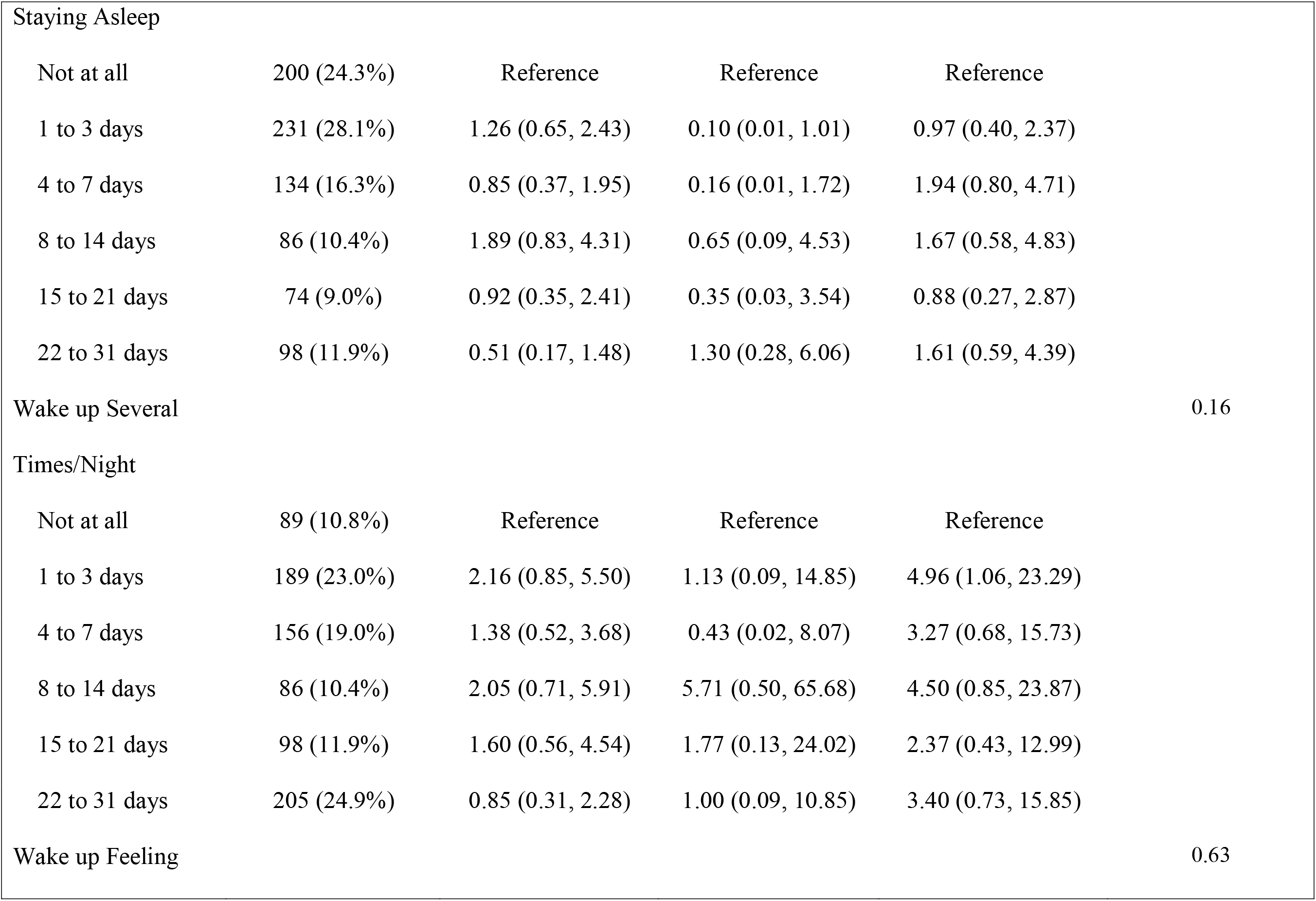
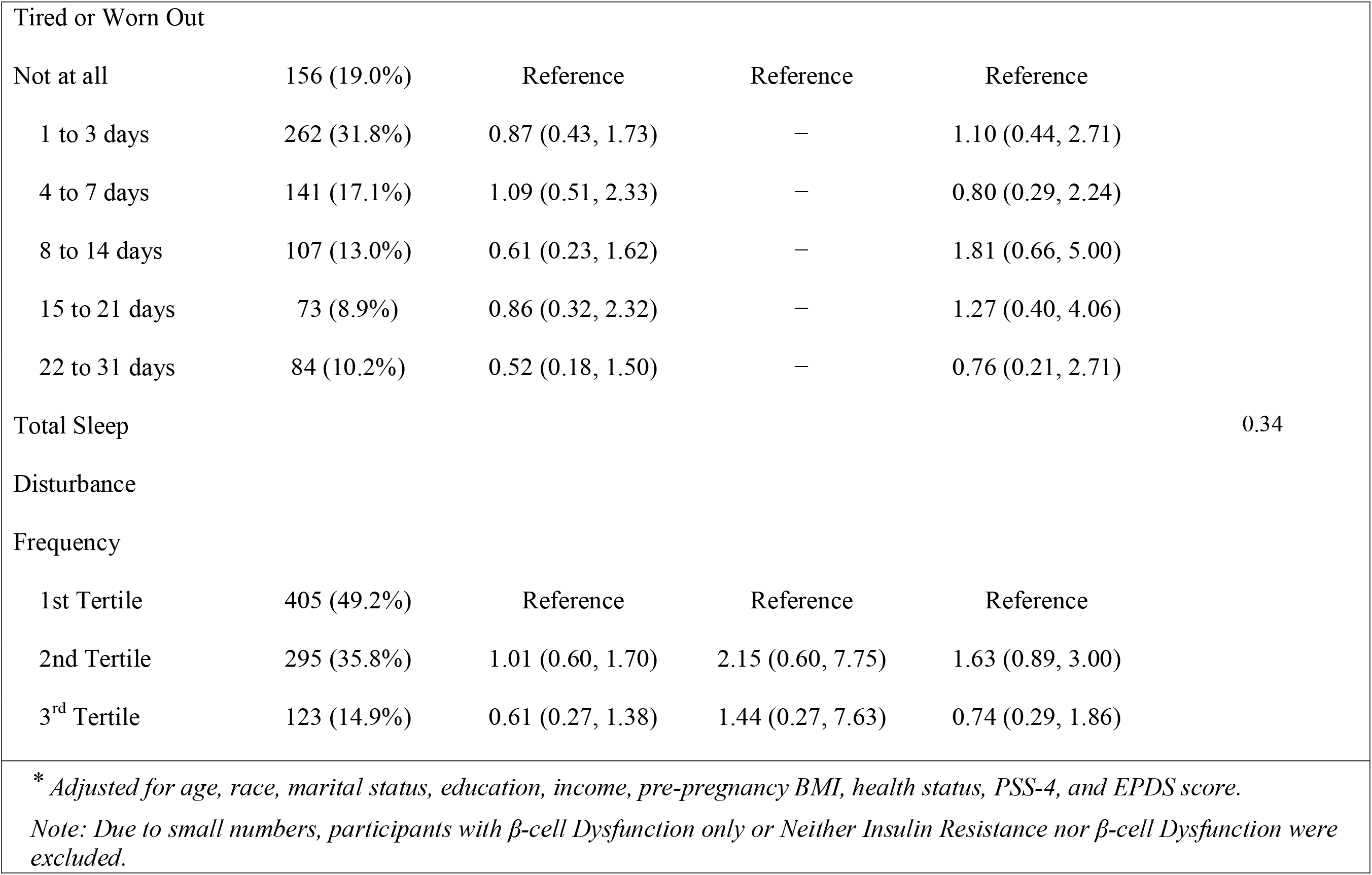
Association between maternal sleep disturbances and GDM subtypes

## Comment

### Principal Findings

This secondary analysis of the GDM2 Trial examined the associations of mid-pregnancy sleep disturbance frequency and mid-pregnancy maternal glycemia and GDM subtypes. We found that the frequency of mid-pregnancy sleep disturbances was not associated with mid-pregnancy maternal glycemia or GDM subtypes.

### Results in the Context of What is Known

Our results differ from other studies that investigated the relationship between other sleep disturbances and maternal glycemia. In a systematic review and meta-analysis of sleep disturbances and perinatal outcomes found that short sleep duration and poor sleep quality were associated with a 1.87 (1.16, 3.01) and 1.53 (1.12, 2.08) odds of GDM, respectively.^22^ Also, among a sample of participants with GDM, short sleep duration was associated with worse glucose control.^10^ We attribute the differences in our findings and findings from prior studies to differences in the sleep disturbances, the timing of sleep assessment, and the period of maternal glycemia. For example, previous studies of sleep disturbances and perinatal outcomes predominantly measured sleep duration, sleep quality, obstructive sleep apnea, or snoring.^22^ In contrast, we measured difficulty falling asleep, staying asleep, waking several times during the night, and waking feeling tired. Sleep duration and sleep quality may be more strongly associated with maternal glycemia during pregnancy than the specific sleep disturbances measured in this study. Also, the timing of sleep assessment may explain this study’s null findings. In the current study, sleep disturbances were assessed between 25 and 32 weeks of gestation. Sleep disturbances likely change in frequency and severity throughout pregnancy, and persistent or worsening sleep disturbances may be more strongly associated with maternal glycemia than an acute experience.

A novel aspect of this study is that we examined associations between sleep disturbances and GDM subtypes, characterized by insulin resistance and/or beta-cell dysfunction.^5, 6^ However, we did not observe differences in the frequency of sleep disturbances by GDM subtypes. This null finding may be explained by the small sample of participants experiencing various GDM subtypes.

### Clinical Implications

Although the results of this study did not support our hypothesis, we addressed an important clinical question. Given the risk for adverse maternal outcomes associated with GDM, examining factors associated with clinical maternal hyperglycemia, insulin resistance, and beta-cell dysfunction or GDM subtypes is important. Our findings from a large study of pregnant women that includes a diverse sample of pregnant women from a large urban hospital in Western Pennsylvania do not support clinicians screening for trouble falling asleep, staying asleep, or waking feeling tired or worn out at the time of GDM screening.

### Research Implications

While we did not find a cross-sectional association between sleep disturbances and maternal glycemia, it is important to note that sleep disturbance frequency likely changes throughout pregnancy. Prior research has shown that sleep changes over the perinatal period is predictive of postpartum depression.^23^ Plausibly, changes in sleep disturbances from early to mid-pregnancy are predictive of maternal hyperglycemia. Future studies should measure patterns of sleep disturbances across pregnancy to better understand its association with maternal glycemia. Likewise, pre-pregnancy exposures contribute maternal risk during pregnancy. Future studies should also measure sleep disturbances pre-pregnancy to understand their independent contribution to maternal hyperglycemia. Also, future studies should measure other sleep health indicators (e.g., sleep regularity and daytime dysfunction) to delineate their relationship between sleep and maternal glycemia. Potentially, measuring a combination of sleep health indicators and disturbances will allow us to identity high risk sleep phenotypes that are more predictive of adverse maternal outcome than individual sleep disturbances.

### Strengths and Limitations

This study has several limitations that deserve brief mention. First, we only measured some commonly occurring sleep disturbances but not other sleep health indicators or disorders. Second, sleep disturbances were measured at only one assessment at the time of GDM screening. Thus, we cannot determine if sleep disturbances in early pregnancy or pre-pregnancy are better predictors of maternal glycemia in pregnancy. Lastly, we used a self-reported measure of sleep disturbances that asked participants to recall their symptom frequency in the past month.

Pregnancy is a period of dynamic change, and we may need repeated measures on sleep disturbances over shorter recall periods (e.g., past week) to identify associations with maternal glycemia. Despite these limitations, a strength of our study was that it leveraged a large RCT of a diverse cohort of women with various maternal glucose levels to examine the relationship between the frequency of sleep disturbances and maternal glycemia.

### Conclusions

In conclusion, sleep disturbance frequency at the time of GDM screening was not associated with maternal glycemia, GDM, or GDM subtype in our cohort. Future research should determine if sleep disturbances measured earlier in pregnancy or in combination with other sleep disturbances have a more meaningful relationship with maternal glycemia.

## Data Availability

All data produced in the present study are available upon reasonable request to the authors

## Clinical Trial Registration Number

Trial Identification Number: NCT02309138 Date of registration: December 5, 2014

## Acknowledgments

None

